# Accuracy of low-density lipoprotein cholesterol estimation equations according to drinking status: A cross-sectional study

**DOI:** 10.64898/2026.10.01.26364472

**Authors:** Jae Yong Lee, Hae Won Jung, Seung-Pyo Hong, Han-Joon Bae

**Author notes:** Corresponding author: Han-Joon Bae, MD., Division of Cardiology, Department of Internal Medicine, Daegu Catholic University Medical Center, 33 Duryugongwon-ro 17-Gil, Nam-gu, Daegu 42472, Republic of Korea.

## Abstract

High low-density lipoprotein cholesterol (LDL-C) and triglyceride (TG) levels are associated with heavy alcohol consumption, but the accuracy of LDL-C estimation equations according to drinking status remains unclear. We analyzed direct LDL-C and drinking-status data from 28,882 participants in the Korea National Health and Nutrition Examination Survey, classified as non/ex-drinkers, moderate drinkers, or binge drinkers. Mean absolute deviation (MAD) was calculated for the Friedewald, Martin-Hopkins, and Sampson equations relative to directly measured LDL-C, and 95% confidence intervals for the median absolute deviation (MeAD) were determined by TG level. Estimation error increased with heavier alcohol consumption and higher TG level. In binge drinkers, all three equations underestimated LDL-C, with the largest deviation at higher TG levels. The Sampson equation showed the lowest MAD and MeAD across drinking categories and TG strata and was most accurate at a TG level of 150–400 mg/dL, whereas the Martin-Hopkins equation was more accurate at TG levels of ≥400 mg/dL. The Sampson or Martin-Hopkins equation estimates LDL-C more accurately than the Friedewald equation regardless of drinking status and should be preferred, with the choice further individualized by TG level in heavy drinkers.

## Introduction

Cardiovascular disease (CVD) remains one of the leading causes of death worldwide, and both its prevalence and associated risk continue to rise [1,2]. Low-density lipoprotein cholesterol (LDL-C) is an independent risk factor for CVD and constitutes one of the primary treatment targets for its prevention [3]. A greater reduction in LDL-C is associated with greater cardiovascular benefit and serves as a benchmark for subsequent treatment goals [3,4].

Numerous factors influence LDL-C level, including diet, exercise, obesity, smoking, and alcohol consumption [3–10]. Alcohol consumption is a major contributor to altered lipid profiles: it disrupts hepatic regulation of cholesterol and triglyceride (TG) metabolism, increases intestinal cholesterol absorption, and accelerates LDL-C production, regardless of the type of alcohol consumed [7,11].

Although direct measurement of LDL-C is the most accurate approach, it is rarely used in clinical practice because it is costly and time-consuming [12,13]. LDL-C is therefore most commonly estimated using the Friedewald equation, which nonetheless has notable limitations. Because individual lipid profiles vary substantially according to drinking status, the Friedewald equation does not account for such variation, and its accuracy becomes particularly uncertain among binge drinkers, who frequently present with markedly elevated TG levels. The Martin-Hopkins and Sampson equations were subsequently developed to provide more accurate LDL-C estimates [14]; however, whether their accuracy likewise varies according to drinking status has not been fully evaluated. We therefore examined the accuracy of these three equations according to drinking status using a large, nationally representative cohort.

## Materials and methods

### Study design and participants

This cross-sectional study used data from the Korea National Health and Nutrition Examination Survey (KNHANES), a nationally representative survey of the Korean population [15]; further details have been described previously [16].

A total of 28,882 Korean adults aged ≥19 years who participated in the KNHANES between January 2009 and December 2023 and had available data on estimated LDL-C and drinking status were included. Participants with TG levels >1,000 mg/dL or missing cholesterol data were excluded. The detailed inclusion and exclusion criteria are shown in S1 Fig.

### Ethics statement

The study was conducted in accordance with the Declaration of Helsinki, and the institutional review board of each participating institution approved the study protocol (IRB No. 2018-01-03-P-A and 2018-01-03-2C-A). KNHANES participants provided written informed consent at the time of the survey.

### Lipid measurement and LDL-C estimation

Blood samples were collected after a minimum 8-hour fast. Lipid concentrations were measured using a homogeneous direct assay (Sekisui Medical Corporation, Tokyo, Japan) on a Hitachi 7600 automated analyzer (Hitachi, Tokyo, Japan), as previously described [16].

### Drinking status

Participants were categorized into three groups according to drinking status. Binge drinkers were defined as adults who consumed 7 or more standard drinks per occasion for men or 5 or more standard drinks per occasion for women. Moderate drinkers were defined as adults who consumed alcohol but did not meet the criteria for binge drinking. Non/ex-drinkers were defined as adults who had never consumed alcohol or had not consumed alcohol within the past year.

Alcohol intake was quantified as the number of standard drinks consumed per drinking session, irrespective of beverage type; one can of beer (355 mL) was considered equivalent to 1.6 standard drinks.

### Statistical analysis

Continuous variables are expressed as medians with interquartile ranges (IQR), and categorical variables are expressed as numbers with percentages. Participants were stratified by TG level as <150, 150–400, and ≥400 mg/dL. Overall precision was defined as the ratio of directly measured LDL-C (dLDL-C) to non-high-density lipoprotein cholesterol (non-HDL-C).

Correlations between dLDL-C and each estimation equation were assessed using scatter plots stratified by drinking status, and concordance was evaluated using the mean absolute error (MAE), the coefficient of determination (R²), and the root mean square error (RMSE). Residual error was defined as the difference between dLDL-C and each estimated LDL-C value. The mean absolute deviation (MAD) and MAE were calculated, and 95% confidence intervals (CIs) for the median absolute deviation (MeAD) were determined for each equation, stratified by dLDL-C level. Pairwise comparisons were adjusted for multiple testing using the Benjamini-Hochberg method. All statistical analyses were performed using R, version 4.1.3 (R Foundation for Statistical Computing, Vienna, Austria).

## Results

### Characteristics according to drinking status

Baseline characteristics of the study population are shown in Table 1 and S1–S3 Tables. The final analysis included 28,882 participants, categorized by drinking status as non/ex-drinkers (n = 2,743), moderate drinkers (n = 11,247), and binge drinkers (n = 14,892). Significant demographic differences were observed across these groups. Binge drinkers were significantly younger (median age, 46.0 years) than non/ex-drinkers (67.0 years; P < 0.001), and the proportion of men was highest among binge drinkers (67.5%) and lowest among non/ex-drinkers (7.5%; P < 0.001). TG level differed significantly according to drinking status (P < 0.001), whereas total cholesterol and high-density lipoprotein cholesterol (HDL-C) levels were similar across groups. Direct LDL-C (dLDL-C) level was higher in moderate drinkers (median, 113.0 mg/dL) than in binge drinkers (111.0 mg/dL; P < 0.001).

**Table 1.**
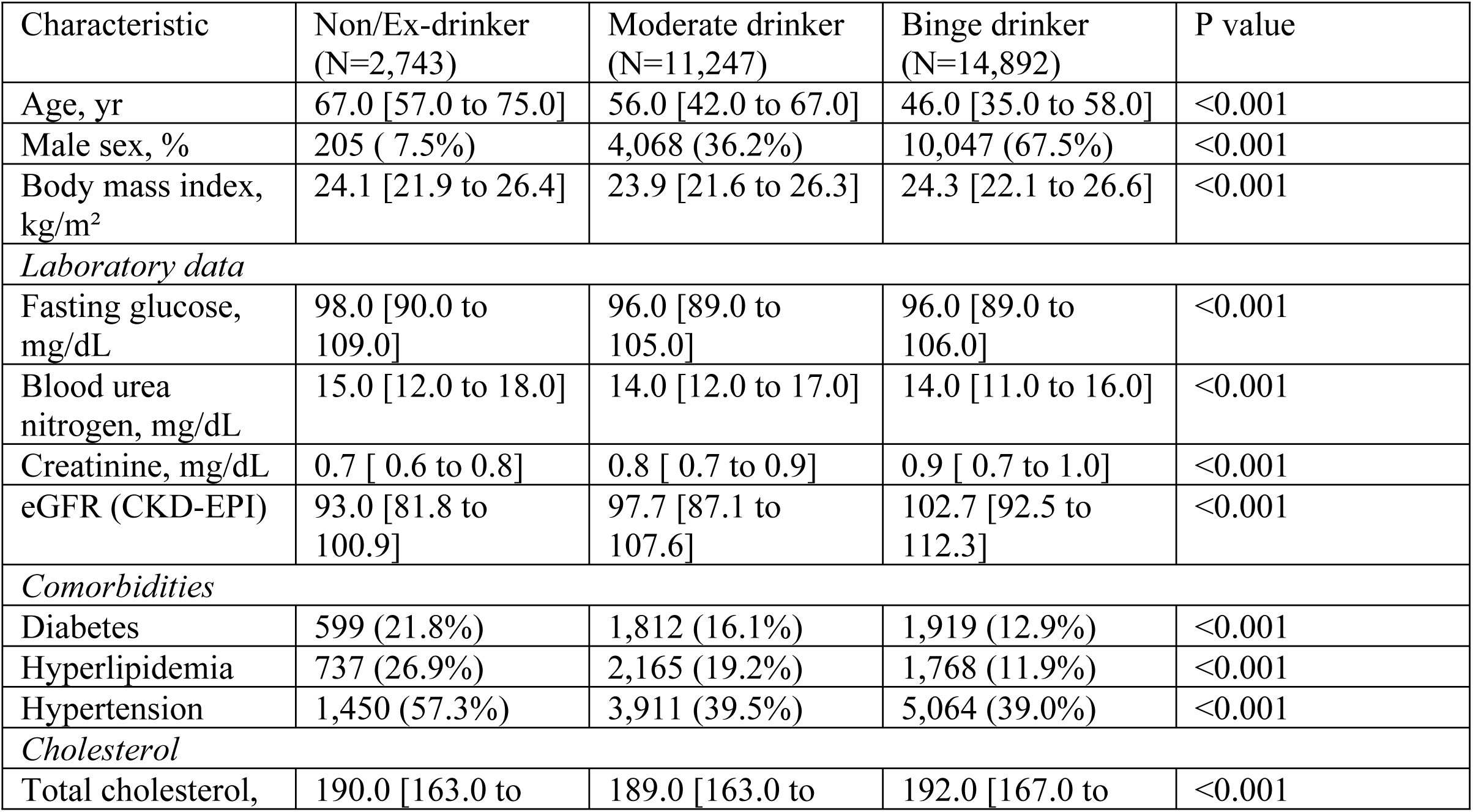
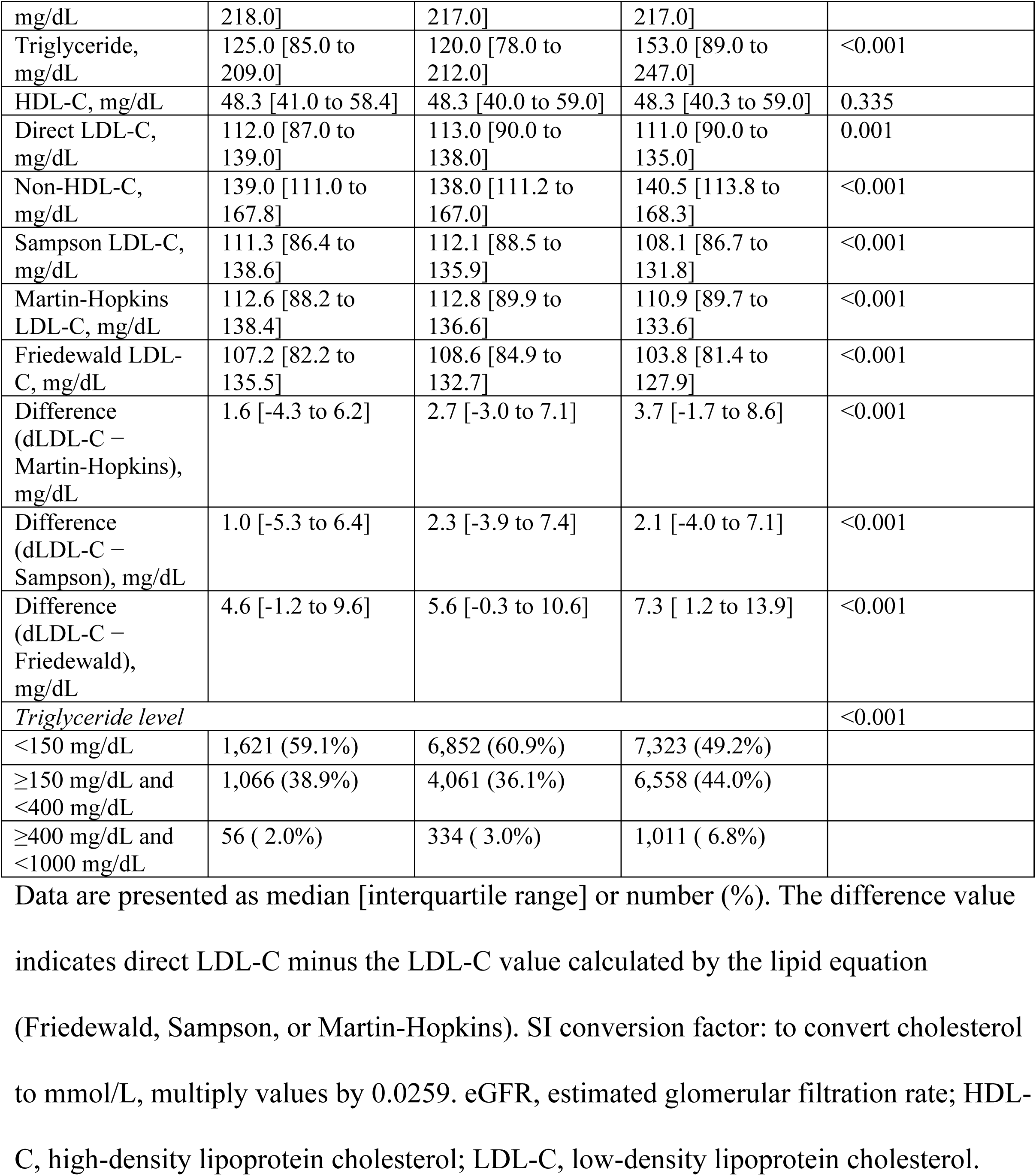
Study population characteristics, overall cohort.

**Table 1.**
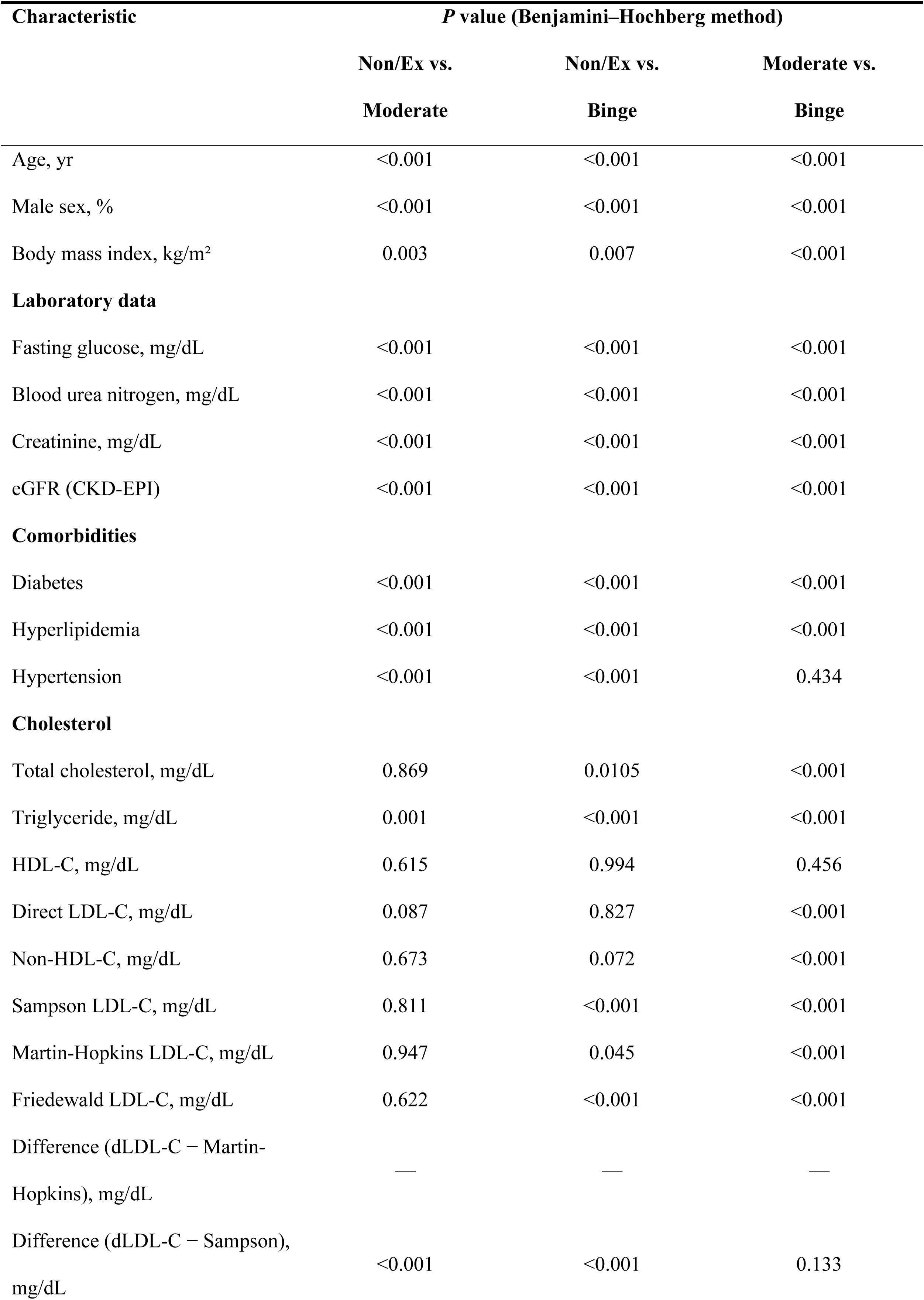
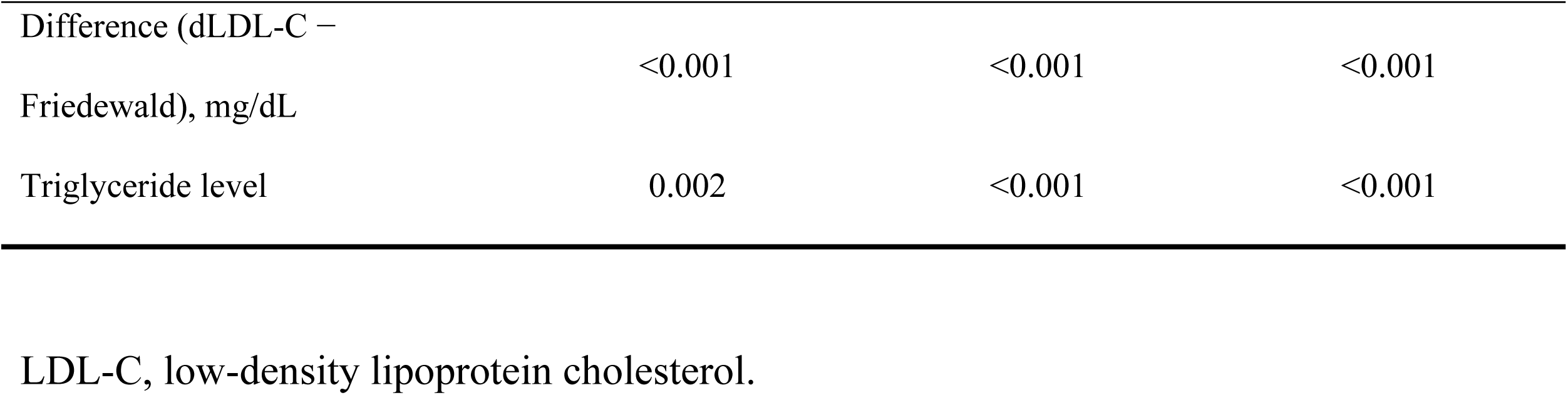
(continued). Pairwise comparisons of the characteristics in Table 1, adjusted for multiple testing (Benjamini-Hochberg method).

### Concordance between direct and estimated LDL-C

We evaluated the concordance between dLDL-C and the values estimated using the Sampson, Martin-Hopkins, and Friedewald equations (Fig 1 and S2 Fig). In binge drinkers, all three equations yielded significantly lower estimates than dLDL-C (P < 0.001 for the Friedewald equation; P = 0.001 for the Sampson and Martin-Hopkins equations). The median estimated LDL-C level in binge drinkers was 108.1 mg/dL using the Sampson equation, 110.9 mg/dL using the Martin-Hopkins equation, and 103.8 mg/dL using the Friedewald equation. After adjustment for multiple comparisons (Benjamini-Hochberg method), the difference values for the Martin-Hopkins and Friedewald equations differed significantly across nearly all pairwise comparisons of drinking status (P < 0.001).

**Fig 1.**
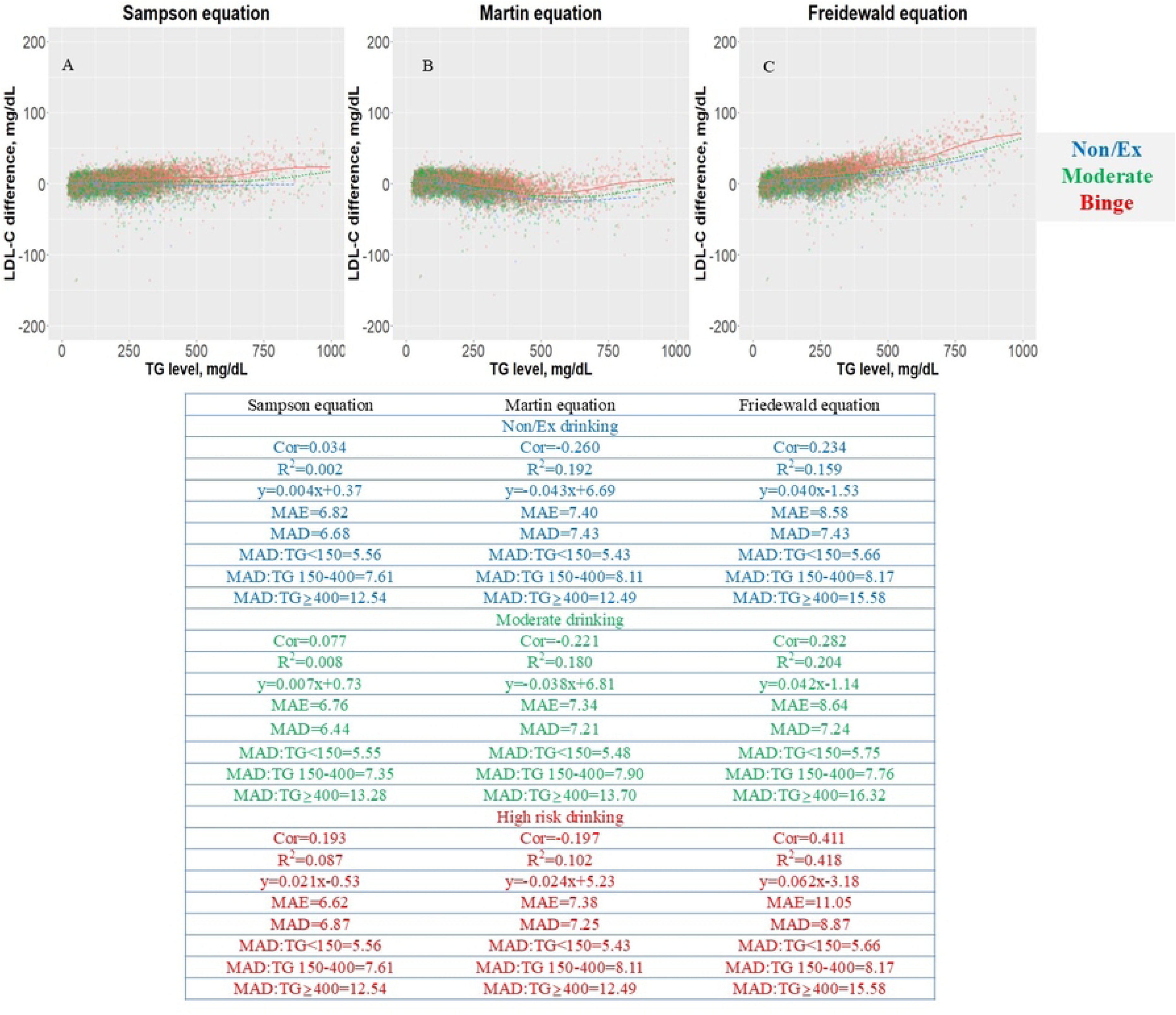
Residual error plots for low-density lipoprotein cholesterol (LDL-C) level by equation, according to drinking status. (A) Difference between the Sampson equation and direct LDL-C (dLDL-C), stratified by triglyceride (TG) level. (B) The Martin-Hopkins equation vs. dLDL-C, stratified by TG level. (C) The Friedewald equation vs. dLDL-C, stratified by TG level. The residual error is the difference between dLDL-C and each estimated LDL-C value. Dots indicate individual samples, colored by drinking status; the solid line indicates the local regression trend. MAD, mean absolute deviation; MAE, mean absolute error; R², coefficient of determination; RMSE, root mean square error.

### Residual error and mean absolute deviation

We further assessed the residual error, defined as the difference between dLDL-C and each estimated LDL-C value, according to drinking status (Fig 1). Across all three drinking groups, the Friedewald equation showed the steepest positive regression slope and the highest coefficient of determination in binge drinkers (y = 0.062x − 3.18, R² = 0.418), indicating that its residual error increased most markedly as LDL-C level rose in this group. In contrast, the Sampson equation showed the weakest correlation between residual error and LDL-C level across all drinking categories (R² = 0.002, 0.008, and 0.087 in non/ex-drinkers, moderate drinkers, and binge drinkers, respectively), reflecting the smallest systematic drift in its estimates. The Martin-Hopkins equation showed an intermediate, negatively sloped relationship (R² = 0.102–0.192). Overall, MAE and MAD followed a similar pattern: the Friedewald equation had the highest MAE in binge drinkers (11.05 mg/dL) compared with the Sampson (6.62 mg/dL) and Martin-Hopkins (7.38 mg/dL) equations, and this discrepancy was most pronounced at a TG level ≥400 mg/dL, where the MAD for the Friedewald equation reached 15.58 mg/dL compared with 12.54 mg/dL for the Sampson equation.

### Mean absolute deviation according to lipid components

The MAD of each equation was further examined according to TG, non-HDL-C, and dLDL-C strata across drinking categories (Fig 2, Table 2, and S4–S6 Tables). In all three drinking groups, the Sampson equation consistently showed the lowest or near-lowest MAD across TG strata, whereas the Friedewald equation showed the highest MAD, and this gap widened progressively with increasing TG level. This pattern was most evident in binge drinkers, in whom the MAD for the Friedewald equation increased sharply at a TG level ≥400 mg/dL (S6 Table), reaching values of 16–30 mg/dL depending on the dLDL-C stratum, whereas the corresponding MAD values for the Sampson and Martin-Hopkins equations remained comparatively lower (10–13 mg/dL). A similar pattern was observed when MAD was stratified by non-HDL-C level, with the largest inter-equation differences again observed in binge drinkers. These findings suggest that the divergence among equations is driven predominantly by TG level and is further amplified by heavier alcohol consumption.

**Fig 2.**
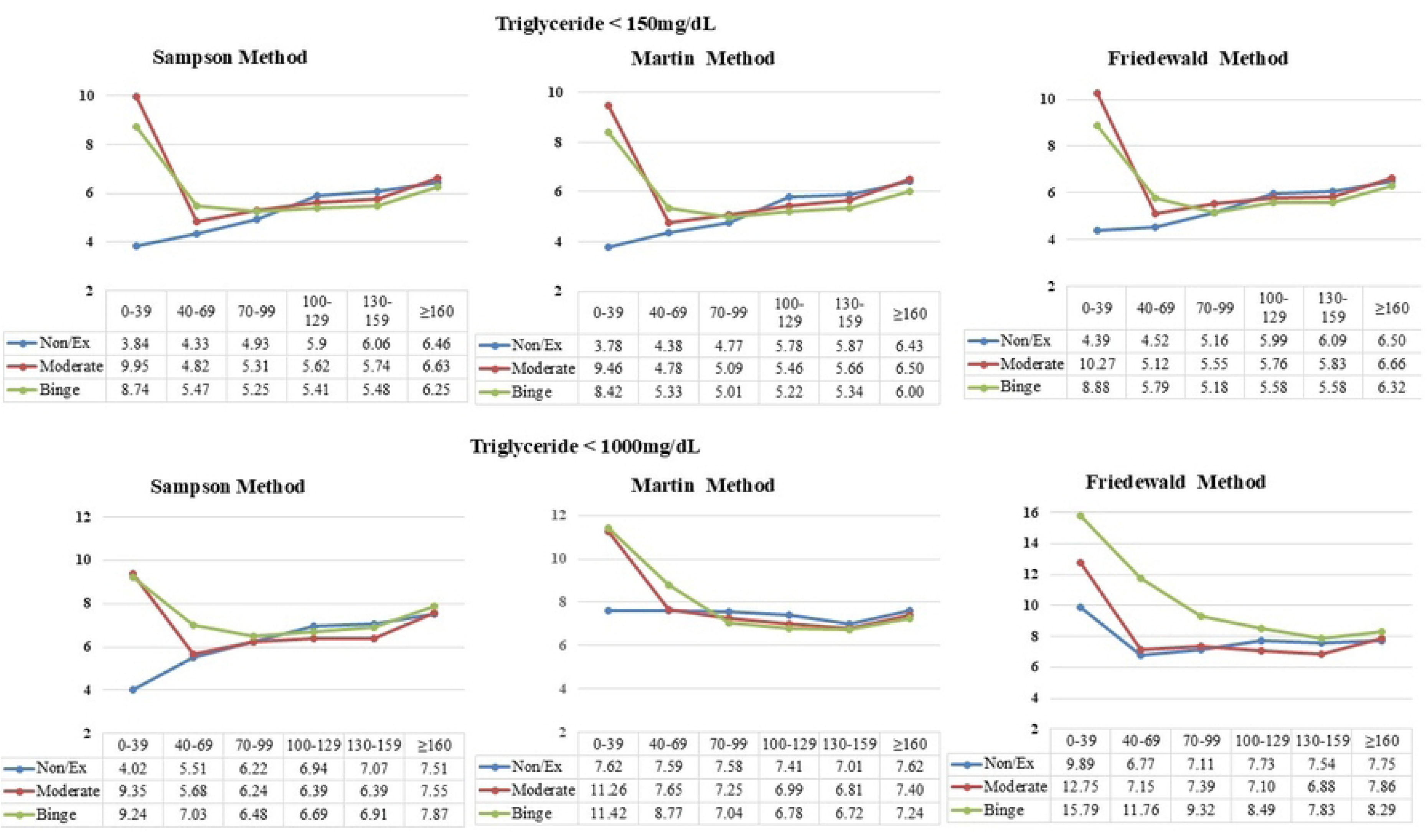
Mean absolute deviation (MAD) of estimated low-density lipoprotein cholesterol (LDL-C), by direct LDL-C (dLDL-C) category, drinking status, and equation. Upper panels: participants with triglyceride (TG) <150 mg/dL. Lower panels: the overall analytic cohort (TG <1,000 mg/dL). Within each panel, MAD is plotted by dLDL-C category (mg/dL) for the Sampson, Martin-Hopkins, and Friedewald equations (left to right), with non/ex-, moderate, and binge drinkers shown as separate lines.

**Table 2.** Mean absolute deviation (MAD) and median absolute deviation (MeAD) with 95% CI of estimated vs. directly measured LDL-C (dLDL-C), overall cohort (TG <1,000 mg/dL).

| dLDL-C category, mg/dL | Sampson |  | Martin-Hopkins |  | Friedewald |  |
| --- | --- | --- | --- | --- | --- | --- |
|  | MAD | MeAD (95% CI) | MAD | MeAD (95% CI) | MAD | MeAD (95% CI) |
| *Non/Ex-drinker* |  |  |  |  |  |  |
| <40 | 4.02 | -2.37 (-8.67 to 3.92) | 7.62 | -2.82 (-9.38 to 3.74) | 9.89 | -2.67 (-9.84 to 4.50) |
| ≥40 to <70 | 5.51 | -1.93 (-3.65 to -0.21) | 7.59 | -1.78 (-3.52 to -0.04) | 6.77 | -2.43 (-4.25 to -0.61) |
| ≥70 to <100 | 6.22 | -0.44 (-1.45 to 0.56) | 7.58 | -0.89 (-1.92 to 0.14) | 7.11 | -0.59 (-1.65 to 0.46) |
| ≥100 to <130 | 6.94 | -2.52 (-3.53 to -1.51) | 7.41 | -2.52 (-3.51 to -1.53) | 7.73 | -3.28 (-4.37 to -2.19) |
| ≥130 to <160 | 7.07 | -2.37 (-3.49 to -1.26) | 7.01 | -1.93 (-3.02 to -0.83) | 7.54 | -2.62 (-3.78 to -1.46) |
| ≥160 | 7.51 | -2.37 (-7.01 to 2.26) | 7.62 | -1.78 (-6.34 to 2.79) | 7.75 | -3.05 (-7.73 to 1.63) |
| *Moderate drinker* |  |  |  |  |  |  |
| <40 | 9.35 | -5.19 (-8.22 to -2.15) | 11.26 | -6.23 (-9.55 to -2.91) | 12.75 | -6.82 (-10.67 to -2.97) |
| ≥40 to <70 | 5.68 | -1.78 (-2.73 to -0.83) | 7.65 | -2.37 (-3.36 to -1.39) | 7.15 | -2.67 (-3.70 to -1.64) |
| ≥70 to <100 | 6.24 | -1.33 (-1.88 to -0.79) | 7.25 | -1.78 (-2.34 to -1.22) | 7.39 | -2.58 (-3.18 to -1.97) |
| ≥100 to <130 | 6.39 | -1.78 (-2.25 to -1.30) | 6.99 | -2.08 (-2.55 to -1.60) | 7.10 | -2.08 (-2.57 to -1.58) |
| ≥130 to <160 | 6.39 | -1.19 (-1.75 to -0.63) | 6.81 | -1.63 (-2.20 to -1.06) | 6.88 | -1.40 (-1.98 to -0.83) |
| ≥160 | 7.55 | -2.45 (-4.75 to -0.14) | 7.40 | -2.30 (-4.59 to -0.01) | 7.86 | -3.00 (-5.33 to -0.67) |
| *Binge drinker* |  |  |  |  |  |  |
| <40 | 9.24 | -1.04 (-4.1 to 2.03) | 11.42 | -5.49 (-9.06 to -1.91) | 15.79 | -10.7 (-15.47 to -5.93) |
| ≥40 to <70 | 7.03 | -2.37 (-3.87 to -0.87) | 8.77 | -2.67 (-4.18 to -1.16) | 11.76 | -6.63 (-8.33 to -4.93) |
| ≥70 to <100 | 6.48 | -1.63 (-2.12 to -1.15) | 7.04 | -1.63 (-2.11 to -1.15) | 9.32 | -3.65 (-4.23 to -3.08) |
| ≥100 to <130 | 6.69 | -1.93 (-2.35 to -1.51) | 6.78 | -1.78 (-2.19 to -1.36) | 8.49 | -2.79 (-3.25 to -2.34) |
| ≥130 to <160 | 6.91 | -1.63 (-2.19 to -1.07) | 6.72 | -1.33 (-1.88 to -0.79) | 7.83 | -2.28 (-2.87 to -1.69) |
| ≥160 | 7.87 | -3.41 (-5.41 to -1.41) | 7.24 | -2.74 (-4.72 to -0.76) | 8.29 | -4.23 (-6.26 to -2.20) |

Data are presented as MAD or MeAD (95% CI), in mg/dL. SI conversion factor: to convert cholesterol to mmol/L, multiply values by 0.0259. CI, confidence interval; dLDL-C, direct low-density lipoprotein cholesterol; MAD, mean absolute deviation; MeAD, median absolute deviation; TG, triglyceride.

### Precision of estimated LDL-C stratified by direct LDL-C level

When precision was stratified directly by dLDL-C level (Table 2), MAD values for all three equations were lowest in the mid-range dLDL-C strata (70–130 mg/dL) and tended to increase at the extremes (dLDL-C <40 mg/dL or ≥160 mg/dL) in all drinking groups. In binge drinkers, the Friedewald equation showed a marked increase in MAD at the lowest dLDL-C stratum (<40 mg/dL; MAD, 15.79 mg/dL) with a MeAD of −10.7 mg/dL (95% CI, −15.47 to −5.93), indicating substantial and significant underestimation of LDL-C in binge drinkers with low dLDL-C. By comparison, the Sampson equation showed the smallest MAD (9.24 mg/dL) and a MeAD closer to zero (−1.04; 95% CI, −4.10 to 2.03) in the same stratum, again supporting the better precision of the Sampson equation at both low and high dLDL-C levels regardless of drinking status.

### Precision of estimated LDL-C according to daily alcohol consumption

We next evaluated the relationship between the residual error of each equation and the amount of alcohol consumed per drinking session, stratified by TG level (Fig 3). In participants with a TG level <150 mg/dL, the residual error showed little association with the amount of alcohol consumed for any equation (R² = 0.002–0.003). However, this association became progressively stronger with increasing TG level. In the TG 150–400 mg/dL group, the Friedewald equation showed the steepest slope (y = 0.364x + 8.06, R² = 0.026) and the largest MeAD (1.31 mg/dL; 95% CI, 0.34 to 2.27), whereas the Sampson and Martin-Hopkins equations showed smaller MeADs (−1.04 and −2.22 mg/dL, respectively). This dose-dependent divergence was most pronounced among participants with a TG level ≥400 mg/dL, in whom the Friedewald equation showed the steepest positive slope (y = 0.892x + 23.81, R² = 0.040) and by far the largest MeAD (7.97 mg/dL; 95% CI, 5.10 to 10.85), indicating that estimation error increased substantially with higher daily alcohol intake among individuals with marked hypertriglyceridemia. In contrast, the Sampson and Martin-Hopkins equations remained comparatively stable across the range of daily alcohol consumption, even at a TG level ≥400 mg/dL (MeAD, −1.63 and −0.89 mg/dL, respectively).

**Fig 3.**
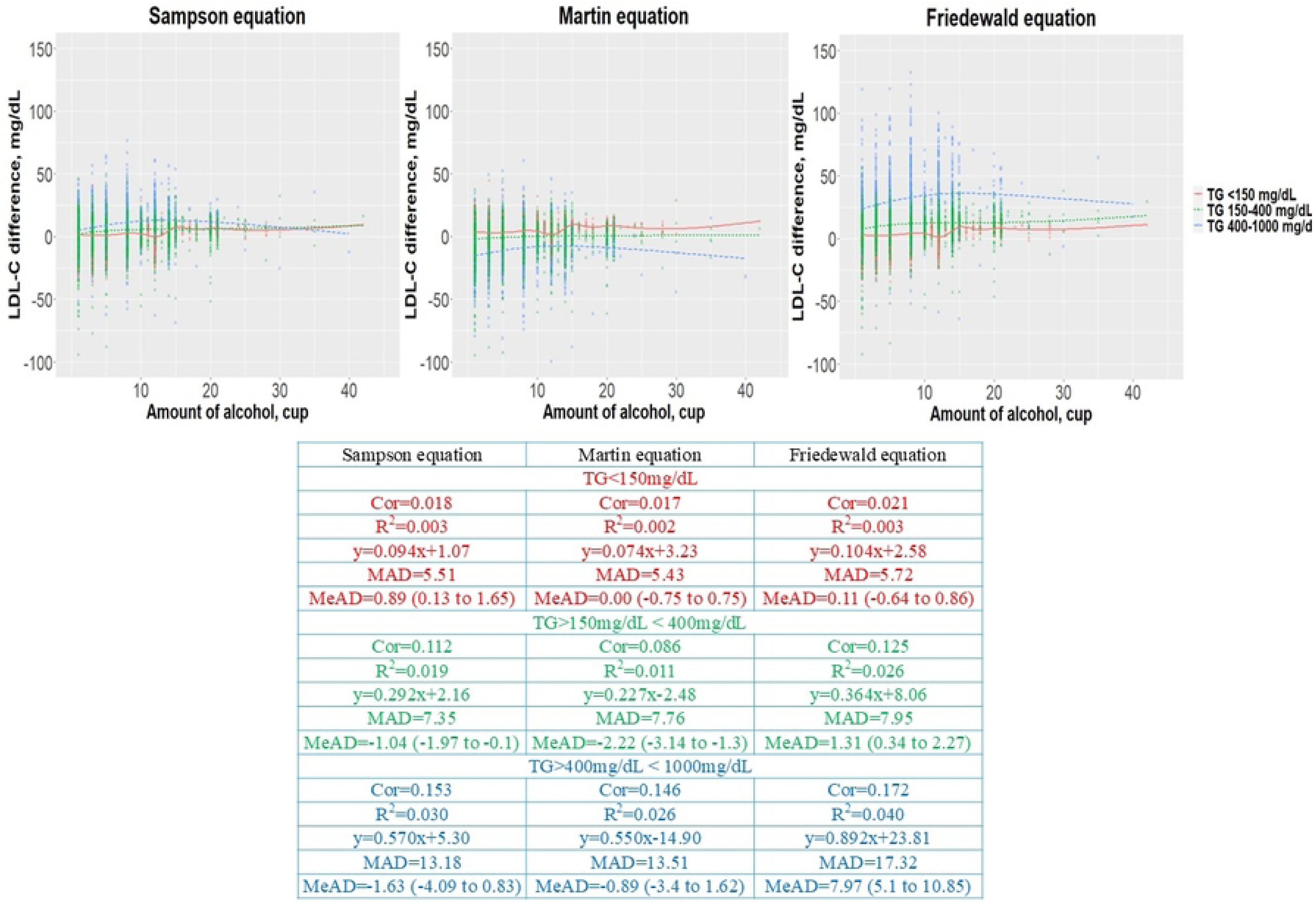
Distribution plots of low-density lipoprotein cholesterol (LDL-C) levels measured by different equations, according to the amount of alcohol consumed per drinking session. (A) Difference between the Sampson equation and direct LDL-C (dLDL-C), stratified by amount of alcohol per day. (B) The Martin-Hopkins equation vs. dLDL-C. (C) The Friedewald equation vs. dLDL-C. Dots indicate individual samples, colored by triglyceride (TG) level; the solid line indicates the local regression trend. MAD, mean absolute deviation; MeAD, median absolute deviation; R², coefficient of determination.

## Discussion

Alcohol consumption is known to alter hepatic and intestinal lipid metabolism, increasing intestinal cholesterol absorption and accelerating hepatic LDL-C production while also elevating TG-rich lipoprotein synthesis [7,11]. A recent large cohort study similarly demonstrated that both increases and decreases in alcohol consumption were associated with corresponding changes in LDL-C and other lipid parameters at repeated health checkups, supporting a direct, modifiable relationship between drinking behavior and lipid profile [17]. Consistent with this mechanism, binge drinkers in our cohort had the youngest age, the highest proportion of men, and the most favorable metabolic and renal profile, yet showed significantly lower dLDL-C (median, 111.0 mg/dL) than moderate drinkers (113.0 mg/dL) despite comparable or higher TG levels; this demographic pattern is consistent with national surveillance data showing that binge and high-risk drinking in Korea is concentrated among younger men [18]. These findings suggest that heavy alcohol intake affects not only the absolute LDL-C level but also its relationship with other lipid components such as TG and non-HDL-C, thereby affecting the accuracy of equation-based LDL-C estimation. All three estimation equations significantly underestimated LDL-C relative to the direct assay in binge drinkers, and this discordance was greater than in the non/ex-drinker and moderate-drinker groups, consistent with previous reports that heavy drinking amplifies TG-related bias in LDL-C calculation [7]. The superior performance of the Sampson and Martin-Hopkins equations over the Friedewald equation observed in this study can be explained by their underlying assumptions. The Friedewald equation estimates very-low-density lipoprotein cholesterol using a fixed TG/5 factor, an assumption that becomes progressively less valid as TG concentration rises, leading to systematic underestimation of LDL-C in individuals with hypertriglyceridemia [12–14]. The Martin-Hopkins equation instead applies an adjustable factor derived from a large reference population stratified by TG and non-HDL-C level, whereas the Sampson equation uses a formula that directly incorporates TG and non-HDL-C without a fixed division factor, allowing both methods to better accommodate the TG elevation frequently observed in heavy drinkers [13,14]. These findings are consistent with several recent validation studies: a large multi-cohort comparison of 24 LDL-C estimation equations against direct assays likewise found that the Martin-Hopkins and Sampson equations outperformed the Friedewald equation, particularly at higher TG levels [19], and a nationwide Korean comparison of newly proposed LDL-C equations similarly reported improved concordance with dLDL-C for non-Friedewald methods [20]. A separate validation study in a Korean population showed that a sex-specific modification of the Sampson-NIH and extended Martin-Hopkins equations improved agreement with directly measured LDL-C beyond the original formulas [21], and an international multicenter validation of the three equations reported similarly superior performance of the Martin-Hopkins and Sampson equations over the Friedewald equation across a wide range of TG levels [22]. Our results extend these principles to the context of drinking status: the Sampson equation showed the most consistent accuracy at a TG level of 150–400 mg/dL across all drinking categories, whereas the Martin-Hopkins equation performed better at a TG level ≥400 mg/dL, a range in which binge drinkers were disproportionately represented. Adjusted pairwise comparisons additionally showed that the difference value for the Sampson equation did not differ significantly between moderate and binge drinkers (P = 0.133), unlike the Martin-Hopkins and Friedewald equations, further supporting the relative robustness of the Sampson equation across drinking intensities.

Beyond categorical drinking status, we examined whether the amount of alcohol consumed per drinking session was associated with estimation error (Fig 3). At normal or mildly elevated TG levels (<150 mg/dL), the residual error of all three equations was largely independent of the amount of alcohol consumed. However, as TG level increased, a clear dose-response relationship emerged, particularly for the Friedewald equation, whose residual error increased most steeply with higher daily alcohol amount among participants with a TG level ≥400 mg/dL (MeAD, 7.97 mg/dL). This dose-dependent pattern parallels recent evidence that graded changes in alcohol intake produce correspondingly graded changes in lipid parameters [17] and indicates that, in individuals who both drink heavily per session and have marked hypertriglyceridemia, reliance on the Friedewald equation could lead to clinically meaningful overestimation of LDL-C accuracy or underestimation of true LDL-C burden. The Sampson and Martin-Hopkins equations were comparatively insensitive to the amount of alcohol consumed even at high TG levels, reinforcing their preferability for cardiovascular risk assessment in individuals with heavy episodic drinking.

This study has several limitations. First, its cross-sectional design precludes causal inference between drinking status and LDL-C estimation accuracy. Second, drinking status and alcohol amount were determined by self-reported questionnaire, which may be subject to recall and social-desirability bias, particularly among binge drinkers, who may underreport their intake. Third, all participants were of Korean ethnicity, so these findings may not be generalizable to other populations with different genetic lipid profiles or drinking patterns. Fourth, participants with TG levels >1,000 mg/dL were excluded, so the comparative accuracy of these equations in severe hypertriglyceridemia related to alcohol use remains uncertain. Finally, information on medication use, such as statins or fibrates, and on the temporal pattern of alcohol consumption relative to blood sampling was not available, and residual confounding by these factors cannot be excluded.

## Conclusions

In this large, nationally representative cross-sectional study, the accuracy of equation-based LDL-C estimation varied significantly according to drinking status and daily alcohol amount, with error increasing progressively with heavier alcohol consumption and higher TG level. The Sampson equation showed the most consistent and precise estimation across drinking categories, particularly at a TG level of 150–400 mg/dL, whereas the Martin-Hopkins equation showed superior accuracy at a TG level ≥400 mg/dL. In contrast, the Friedewald equation showed the greatest systematic error, especially among binge drinkers with elevated TG levels. Therefore, the Sampson or Martin-Hopkins equation, rather than the traditional Friedewald equation, should be considered the primary tool for estimating LDL-C regardless of drinking status, with the choice of equation further individualized according to TG level in heavy drinkers.

## Data Availability

The data underlying this study are publicly available from the Korea National Health and Nutrition Examination Survey (KNHANES) website (https://knhanes.kdca.go.kr/knhanes/main.do) after registration. The authors had no special access privileges that others would not have.

## Acknowledgments

We thank the Korean Ministry of Health and Welfare for providing the data used in this study. A large language model was used to improve the English language of the manuscript; the authors reviewed and edited all output and take full responsibility for the content. This work was supported by the grant of Research Institute of Medical Science, Daegu Catholic University(2022)

## Supporting information

**S1 Fig. Flow diagram of participant selection.** Inclusion and exclusion criteria for participants from the Korea National Health and Nutrition Examination Survey (KNHANES), 2009–2023.

**S2 Fig. Correlation of estimated vs. direct low-density lipoprotein cholesterol (LDL-C) levels according to drinking status.**

**S1 Table. Study population characteristics for triglyceride levels of <150 mg/dL.**

**S2 Table. Study population characteristics for triglyceride levels of ≥150 and <400 mg/dL.**

**S3 Table. Study population characteristics for triglyceride levels of ≥400 and <1,000 mg/dL.**

**S4 Table. Mean absolute deviation (MAD) and median absolute deviation (MeAD) with 95% CI of measured direct low-density lipoprotein cholesterol (dLDL-C) vs. estimated LDL-C in patients with triglyceride levels of <150 mg/dL.**

**S5 Table. Mean absolute deviation (MAD) and median absolute deviation (MeAD) with 95% CI of measured direct low-density lipoprotein cholesterol (dLDL-C) vs. estimated LDL-C in patients with triglyceride levels of ≥150 and <400 mg/dL.**

**S6 Table. Mean absolute deviation (MAD) and median absolute deviation (MeAD) with 95% CI of measured direct low-density lipoprotein cholesterol (dLDL-C) vs. estimated LDL-C in patients with triglyceride levels of ≥400 and <1,000 mg/dL.**

## Notes

### Competing Interest Statement

The authors have declared no competing interest.

### Author Declarations

KNHANES participants provided written informed consent at the time of the survey.

